# Universal Opt-Out Hepatitis C Virus Testing and Treatment on Entry in California State Prisons

**DOI:** 10.64898/2026.03.20.26348733

**Authors:** Ziping Ye, Kim Lucas, Nathan Furukawa, Amanda Honeycutt, Donna Kalauokalani, Amy Krawiec, Teresa Puente, Joshua A. Salomon, Marissa B. Reitsma

## Abstract

**Background:** Correctional facilities are vital venues for expanding testing and treatment for hepatitis C virus (HCV) infections, essential components of national hepatitis C elimination plans.

**Objective:** This study characterizes HCV testing and treatment outcomes among individuals entering incarceration into California state prisons, overall, by year, and by key individual-level characteristics.

**Methods:** We analyzed individual-level electronic health record data from all adults entering California prisons (“entrants”) between July 1, 2016 and June 30, 2023. We quantified the percentages of entrants receiving an HCV antibody test within four weeks of entry, the percentage antibody positive among tested, the percentage RNA positive among antibody positive, and the percentage initiating direct acting antiviral (DAA) treatment within one year among RNA positive.

**Results:** Of entrants, 133,639 (76%) were tested for HCV antibody, 25,455 (19% of tested) were ever HCV-infected, and 16,738 (66% of ever infected) were currently infected. Among individuals currently infected, 7,479 (45%) initiated DAA treatment within one year. Individuals with identified SUD had 3.2 times higher antibody positivity and 1.3 times higher proportions initiating DAA, compared to individuals not having an identified SUD.

**Discussion:** We show that HCV testing and treatment in California prisons, a central component of national hepatitis C elimination efforts, supported effective and equitable increases in access to hepatitis C treatment, particularly for those with SUD.

**Disclaimer:** The findings and conclusions in this report are those of the authors and do not necessarily represent the official position of the Centers for Disease Control and Prevention or other authors’ affiliated institutions.

## Background

Correctional facilities are vital venues for expanding testing and treatment for hepatitis C virus (HCV) infections, essential components of national hepatitis C elimination plans.^1,2^ Compared to a general population prevalence of current HCV infection of 1.6% in the United States,^3^ a 2023 study estimated a mean prevalence of 8.7% (range 1.4% to 24.5%) across state prison systems.^4^ In July 2016, California Correctional Health Care Services (CCHCS) began implementing multiple strategies toward achieving elimination of hepatitis C within the state prison system, including universal opt-out testing on entry, gradual treatment eligibility expansions, and linkage to substance use disorder (SUD) treatment.^5^ This study characterizes HCV testing and treatment outcomes among individuals entering incarceration into California state prisons, overall, by year, and by key individual-level characteristics.

## Methods

We analyzed individual-level electronic health record data from all adults entering California prisons (“entrants”) between July 1, 2016 and June 30, 2023, who had at least six months of follow-up data while incarcerated. We quantified the percentages of entrants receiving an HCV antibody test within four weeks of entry, the percentage antibody positive (ever infected) among tested, the percentage RNA positive (currently infected) among antibody positive, and the percentage initiating direct acting antiviral (DAA) treatment within one year among RNA positive. To assess policy implementation, we report on outcomes overall, and by year. We also report outcomes stratified by sex, age group, race/ethnicity, prior incarceration in a California prison, anticipated length of incarceration, and SUD history. Additional details are provided in the Supplement.

## Results

During July 2016 through June 2023, 176,967 individuals meeting eligibility criteria entered California prisons. Of entrants, 133,639 (76%) were tested for HCV antibody, 25,455 (19% of tested) were ever HCV-infected, and 16,738 (66% of ever infected) were currently infected (Table 1). Among individuals currently infected, 7,479 (45%) initiated DAA treatment within one year. Among individuals who initiated DAA treatment within one year, 51% did so within six months of their positive test at entry. Among individuals who did not initiate DAA treatment, 46% were released between six months and one year of entry. From fiscal year 2017-2018 to 2022-2023, testing and treatment percentages increased (80% to 92% and 20% to 76%, respectively), antibody positivity remained largely stable (18% to 20%), and RNA positivity decreased (72% to 55%) (Figure 1).

**Table 1.** Hepatitis C Virus Testing and Treatment on Entry to California State Prisons by Demographic and Carceral Characteristics, Fiscal Year 2016-2017 to 2022-2023.

|  | Number Entering | Antibody Tests (%) | Positive Antibody Tests (%) | Positive RNA Tests (%) | DAA Initiations (%) |
| --- | --- | --- | --- | --- | --- |
| **Overall** |  |  |  |  |  |
| Overall | 176,967 | 133,639 (75.5) | 25,455 (19.0) | 16,738 (65.8) | 7,479 (44.7) |
| **Sex** |  |  |  |  |  |
| Female | 10,631 | 10,041 (94.5) | 1,413 (14.1) | 786 (55.6) | 267 (34.0) |
| Male | 166,336 | 123,598 (74.3) | 24,042 (19.5) | 15,952 (66.4) | 7,212 (45.2) |
| **Age Group<sup>1</sup>** |  |  |  |  |  |
| 18-29 | 64,047 | 48,049 (75.0) | 6,096 (12.7) | 4,487 (73.6) | 1,951 (43.5) |
| 30-49 | 91,852 | 69,858 (76.1) | 14,270 (20.4) | 9,504 (66.6) | 4,359 (45.9) |
| 50+ | 21,068 | 15,732 (74.7) | 5,089 (32.3) | 2,747 (54.0) | 1,169 (42.6) |
| **Race/Ethnicity<sup>2</sup>** |  |  |  |  |  |
| Hispanic | 82,359 | 61,615 (74.8) | 12,145 (19.7) | 8,487 (69.9) | 3,915 (46.1) |
| Non-Hispanic American Indian/Alaska Native | 1,906 | 1,531 (80.3) | 472 (30.8) | 263 (55.7) | 110 (41.8) |
| Non-Hispanic Asian or Pacific Islander | 2,620 | 2,027 (77.4) | 97 (4.8) | 52 (53.6) | 23 (44.2) |
| Non-Hispanic Black | 40,996 | 30,503 (74.4) | 1,760 (5.8) | 1,038 (59.0) | 447 (43.1) |
| Non-Hispanic White | 42,511 | 33,137 (77.9) | 10,291 (31.1) | 6,422 (62.4) | 2,764 (43.0) |
| Non-Hispanic Other | 6,575 | 4,826 (73.4) | 690 (14.3) | 476 (69.0) | 220 (46.2) |
| **Previously Incarcerated in a California State Prison** |  |  |  |  |  |
| Yes | 102,929 | 73,287 (71.2) | 20,790 (28.4) | 13,611 (65.5) | 6,207 (45.6) |
| No | 74,038 | 60,352 (81.5) | 4,665 (7.7) | 3,127 (67.0) | 1,272 (40.7) |
| **Years Expected to Serve<sup>3</sup>** |  |  |  |  |  |
| Less than One Year | 60,809 | 48,288 (79.4) | 9,085 (18.8) | 5,876 (64.7) | 1,677 (28.5) |
| One or Two Years | 71,218 | 52,971 (74.4) | 11,002 (20.8) | 7,222 (65.6) | 3,895 (53.9) |
| Three to Nine Years | 31,745 | 22,314 (70.3) | 3,894 (17.5) | 2,647 (68.0) | 1,421 (53.7) |
| Ten or More Years | 13,195 | 10,066 (76.3) | 1,474 (14.6) | 993 (67.4) | 486 (48.9) |
| **SUD Status<sup>4</sup>** |  |  |  |  |  |
| Assessed, SUD identified | 45,593 | 39,891 (87.5) | 12,814 (32.1) | 8,354 (65.2) | 4,836 (57.9) |
| Screened Negative or Assessed, no SUD identified | 60,943 | 50,440 (82.8) | 5,032 (10.0) | 3,119 (62.0) | 1,374 (44.1) |
| Not Screened or Assessed (No SUD Program) | 70,431 | 43,308 (61.5) | 7,609 (17.6) | 5,265 (69.2) | 1,269 (24.1) |
| **Fiscal Year<sup>5</sup>** |  |  |  |  |  |
| 2016 - 2017 | 31,328 | 4,995 (15.9) | 829 (16.6) | 476 (57.4) | 26 (5.5) |
| 2017 - 2018 | 32,531 | 26,024 (80.0) | 4,628 (17.8) | 3,327 (71.9) | 675 (20.3) |
| 2018 - 2019 | 32,674 | 28,541 (87.4) | 5,483 (19.2) | 3,963 (72.3) | 1,716 (43.3) |
| 2019 - 2020 | 20,845 | 18,568 (89.1) | 3,463 (18.7) | 2,426 (70.1) | 752 (31.0) |
| 2020 - 2021 | 13,467 | 12,165 (90.3) | 2,348 (19.3) | 1,460 (62.2) | 845 (57.9) |
| 2021 - 2022 | 22,788 | 21,978 (96.4) | 4,494 (20.4) | 2,760 (61.4) | 1,695 (61.4) |
| 2022 - 2023 | 23,334 | 21,368 (91.6) | 4,210 (19.7) | 2,326 (55.2) | 1,770 (76.1) |

Abbreviations: Substance Use Disorder (SUD); Direct-Acting Antiviral (DAA)
<sup>1</sup>Age group is defined at entry and is reported in years.
<sup>2</sup>Race/ethnicity groups are mutually exclusive. Individuals documented to be part of more than one race/ethnicity group were included in the 'Non-Hispanic Other' group.
<sup>3</sup>Refers to the expected or observed sentence duration.
<sup>4</sup>Routine SUD screening and assessment began in January 2020 (99.7% of individuals 'Not Screened or Assessed' entered prior to January 2020). Individuals were considered to have been identified with SUD if A) their NIDA-Modified ASSIST Substance Involvement Score indicated Moderate Risk or High Risk (score of 4+) for any substance category (cannabis, cocaine, prescription stimulants, methamphetamine, inhalants, sedatives, hallucinogens, street opioids, prescription opioids), B) they had ever been treated with a medication for opioid use disorder, or C) they met American Society of Addiction Medicine Level of Care criteria for opioid treatment services. Individuals were considered to have no SUD identified if they screened negative on the NIDA Quick Screen or were not identified with SUD using any of the aforementioned criteria.
<sup>5</sup>Fiscal year spanning July to June.

**Figure 1.**
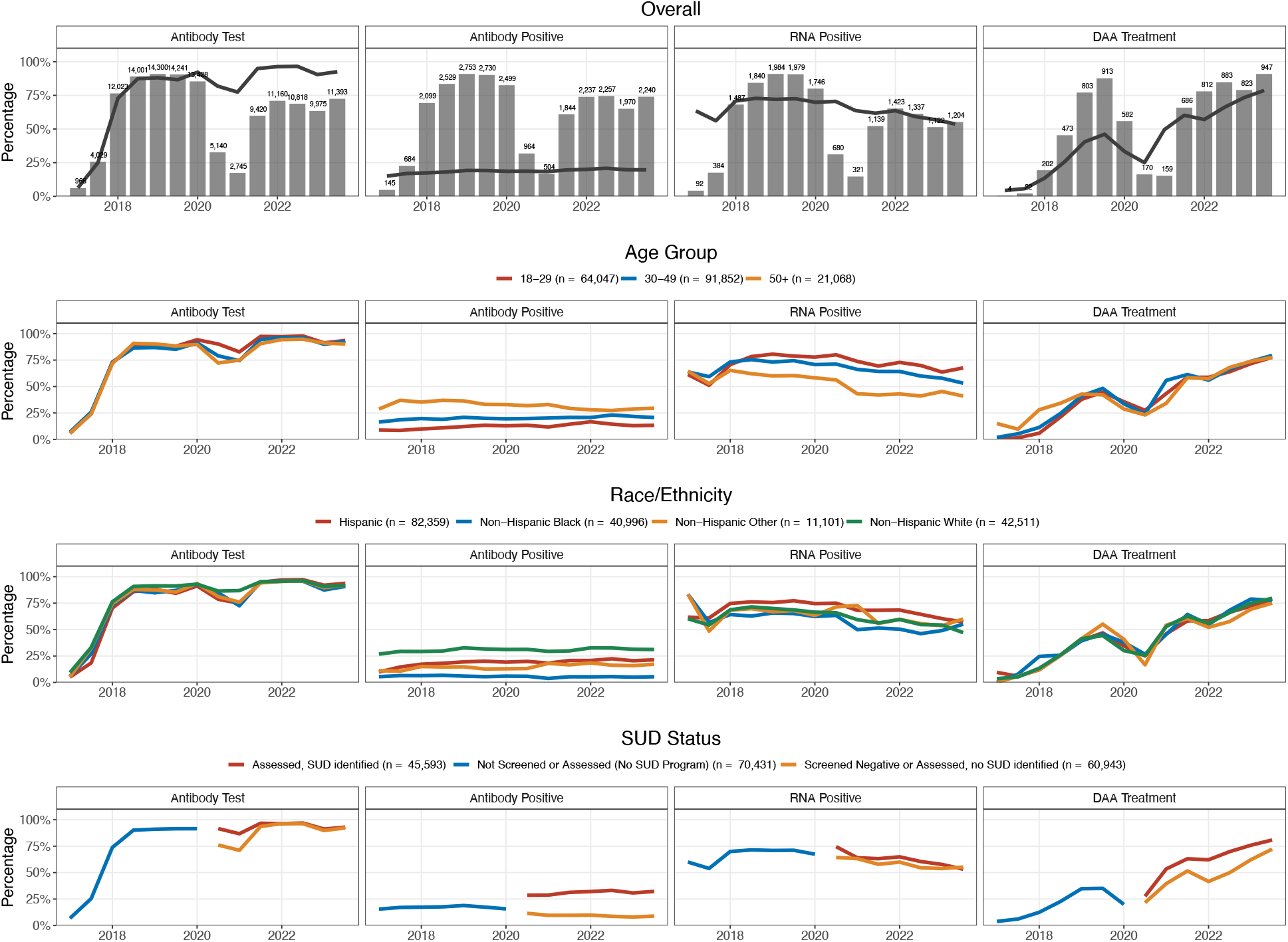
Changes in the Hepatitis C Virus Testing and Treatment on Entry to California State Prisons, Overall and by Age, Race/Ethnicity, and Substance Use Disorder Status, Fiscal Year 2016-2017 to 2022-2023. Note: The decline in antibody testing and DAA treatment observed in 2020 and 2021 coincide with the COVID-19 pandemic, which considerably impacted the prison health system. Race/ethnicity groups are mutually exclusive. Individuals documented to be part of more than one race/ethnicity group were included in the ‘Non-Hispanic Other’ group. In the figure, “Non-Hispanic Other” race/ethnicity also includes “Non-Hispanic American Indian/Alaska Native” and “Non-Hispanic Asian or Pacific Islander” groups due to small numbers. Individuals were considered to have been identified with SUD if A) their NIDA-Modified ASSIST Substance Involvement Score indicated Moderate Risk or High Risk (score of 4+) for any substance category (cannabis, cocaine, prescription stimulants, methamphetamine, inhalants, sedatives, hallucinogens, street opioids, prescription opioids), B) they had ever been treated with a medication for opioid use disorder, or C) they met American Society of Addition Medicine Level of Care criteria for opioid treatment services. Individuals were considered to have no SUD identified if they screened negative on the NIDA Quick Screen or were not identified with SUD using any of the aforementioned criteria. Age group categories are reported in years. Lines connect values calculated over six-month periods. The gaps in lines on the SUD Status panels separates the last period before SUD program activation and the first period after SUD program activation. SUD: Substance Use Disorder. DAA: Direct-Acting Antiviral.

Although antibody and RNA positivity percentages differed across age and racial/ethnic groups, access to HCV testing and treatment was similar. The prevalence of current HCV infection among tested entrants was highest among male, older, and Hispanic entrants, as well as entrants who had previously been incarcerated in a California prison. Individuals with identified SUD had 3.2 times higher antibody positivity and 1.3 times higher proportions initiating DAA, compared to individuals not having an identified SUD (Table 1).

## Discussion

CCHCS implemented universal opt-out HCV testing and treatment on entry, as part of a wide-scale effort to eliminate hepatitis C, achieving largely equal access to care across demographic groups. Although we document subgroup heterogeneity in RNA positivity, the high overall current infection rate among entrants (13%) supports universal screening strategies.^3,6^ Higher DAA initiation among RNA positive individuals identified with SUD underscores the value of integrated SUD and hepatitis C treatment programs. Future work should evaluate the long-term health and economic effects of testing and treatment strategies. Our primary limitation is that results from California may not generalize to other states due to differences in prison systems, demographics, and epidemiology. We show that HCV testing and treatment in California prisons, a central component of national hepatitis C elimination efforts, supported effective and equitable increases in access to hepatitis C treatment, particularly for those with SUD.

## Data Availability

Individual-level data from California Correctional Health Care Services are not publicly accessible and require a data use agreement. Analytic code and aggregate results are accessible at: https://github.com/PPML/cchcs_hcv_screening_manuscript.

## Ethics Statement

This study was determined by the Stanford University Institutional Review Board to be non–human subjects research (Protocol #72584).

## Exhibits

