## Supplement for "Universal Opt-Out Hepatitis C Virus Testing and Treatment on Entry in California State Prisons"

### Eligibility

Our analytic cohort captured all adult individuals (ages 18 years and older) entering or re-entering the California state prison system at any point during July 1, 2016 through June 30, 2023 with at least six months of follow-up data.

### Data Processing

#### *Demographic and Carceral Characteristics*

We extracted data on demographic and carceral characteristics for individuals in our analytic cohorts. Demographic characteristics included documented sex (male, female), age group (18-29, 30-49, 50+ years), and race/ethnicity (Hispanic, non-Hispanic American Indian/Alaska Native, non-Hispanic Asian or Pacific Islander, non-Hispanic Black, non-Hispanic White, non-Hispanic Other). Carceral characteristics included whether an individual was previously incarcerated in the California state prison system (yes, no) and their sentence length (less than one year, one to two years, three to nine years, ten or more years).

Age was defined based on documented birth year at entry. Race/ethnicity groups are mutually exclusive. The Hispanic group included individuals of any race. Individuals with race/ethnicity documented as unknown or with multiple documented racial/ethnic groups were included in the non-Hispanic Other racial/ethnic group.

Sentence length was defined as the time between entry and anticipated release. For current residents at the end of our data follow-up period, we used estimated release week to define sentence length. For current residents without an estimated release week, we assumed sentence length was 10+ years to reflect a lifetime sentence.

#### *Hepatitis C Virus Testing*

California Correctional Health Care Services hepatitis C virus (HCV) laboratory generally uses HCV antibody and reflexed RNA tests. In some rare cases where prior HCV infection is documented, only RNA testing is performed. When constructing the HCV care cascade, we assumed a positive antibody test if an RNA test was performed without an antibody test. Since dates were aggregated to weekly indicators to protect privacy, if multiple tests performed in the same week yielded different results, we retained the positive test only.

#### *Direct Acting Antiviral Treatment*

We only included direct-acting antiviral (DAA) treatments for HCV in our analysis. As a result, a small number of individuals who were treated with pre-DAA regimens (e.g., pegylated interferon and ribavirin) were considered untreated in the present analysis. We used a one-year follow-up from the date of entry to identify DAA initiation.

*Substance Use Disorder (SUD) Assessment*

Individuals were considered to have been identified with SUD if A) their NIDA-Modified ASSIST (https://nida.nih.gov/sites/default/files/pdf/nmassist.pdf) Substance Involvement Score indicated Moderate Risk or High Risk (score of 4+) for any substance category (cannabis, cocaine, prescription stimulants, methamphetamine, inhalants, sedatives, hallucinogens, street opioids, prescription opioids), B) they had ever been treated with a medication for opioid use disorder, or C) they met American Society of Addition Medicine Level of Care criteria (https://downloads.asam.org/sitefinity-production-blobs/docs/default-source/quality-science/asam_loc-assessment-guide-print_all.pdf) for opioid treatment services. Individuals were considered to have no SUD identified if they screened negative on the NIDA Quick Screen or were not identified with SUD using any of the aforementioned criteria. SUD screening and identification were defined using data from full incarceration periods (defined by entry and exit dates). Consistency within incarceration periods was implemented by applying the following hierarchy: identified with SUD, identified as not having SUD, not screened or assessed.
